# Governing birth in a pandemic: A policy analysis of maternity hospital restrictions in Ontario during the COVID-19 pandemic

**DOI:** 10.1101/2025.09.17.25336025

**Authors:** Maria Ahmed

## Abstract

At the beginning of the COVID-19 pandemic, hospitals around the world modified policies for many services, including maternity care, to limit viral transmission. While early decisions were made under significant uncertainty, it is not clear how much consistency there was across jurisdictions, or whether some of these policies inadvertently and disproportionately impacted maternal and infant health for marginalized populations. Using a digital archive database, this study examines how maternity policies evolved during the COVID-19 pandemic across Ontario hospitals, assesses equity implications for marginalized communities, and evaluates the extent to which policy changes aligned with data-driven public health risk assessments. A thematic content analysis of obstetric policy documents on 13 Ontario hospital websites between 2020 and 2023 explores three policy areas with equity implications for maternal care: visitor access, support partner restrictions, and doula care limitations. Using a health equity perspective, findings show a high degree of variability in how the Ontario Ministry of Health policies were implemented both within and across hospitals and raise concerns about equity for marginalized populations, social justice in health, and evidence-informed policy alignment.

## Introduction

In Ontario, much like other parts of the world, institutions modified policies with respect to maternity services in response to the COVID-19 pandemic. While decisions were made under uncertainty and with the utilitarian purpose of limiting viral transmissions, many policies placed restrictions on fundamental caregiving practices by separating mothers and infants in both confirmed and unconfirmed cases of infection and excluding support persons for the labouring mother irrespective of infection status [1,2]. Moreover, these policies were implemented inconsistently and with a high degree of variability across jurisdictions [3,4]. Research suggests such policies may have negatively impacted the mental health of both birthing mothers and their caregivers, with intergenerational consequences reflected in lower levels of exclusive and overall breastfeeding among women who reported poor pandemic birth experiences [5].

Balancing the health care needs of a minority population and the needs of the broader public is inherently challenging and requires careful deliberation on what constitutes “essential care” for birthing people. Early in the COVID-19 pandemic, some jurisdictions grappled with this definition by examining the extent of support permitted (none, one person, or more), the timing of need (prenatal, labour, postpartum), and the conditions for support (asymptomatic individuals, vaccinated partners) [6]. Jurisdictions adopted varying combinations of these parameters [4], suggesting that policy decisions were guided more by infection control imperatives or bureaucratic considerations than by a coherent understanding of essential care in maternal health. Moreover, early guidelines for labour and delivery patients during the COVID-19 pandemic more closely followed the objective of reducing risk of viral spread by minimizing direct contact with hospitals and isolating “the COVID-19–positive woman” [7]. This raises critical questions of which factors were prioritized in the development and implementation of public health measures for maternal health and what were the implications on marginalized populations.

While scholars have emphasized the importance of incorporating social justice and equity considerations in the implementation of public health measures [8,9], the early pandemic response largely emphasized COVID-19 as the “great equalizer”, thereby removing equity from discussion in policy development and implementation [10,11]. Moreover, data was often framed as a neutral, objective tool for decision-making and action [12], yet this approach risked overlooking the lived realities of those most impacted, as evidenced by the strong public backlash to restrictive maternity policies such as “birthing alone” [13,14]. The high degree of policy variability may also have led some women to opt out of hospital births altogether [15,16] while eroding trust in public health policies for others [3]. This may have long-term implications on compliance levels for future health guidance.

To better understand the context of hospital-level policymaking, this study has three key objectives:

1. To examine how maternal care policies evolved during the COVID-19 pandemic in hospitals across Ontario.
2. To evaluate the health equity impacts of these policies on marginalized populations.
3. To assess the extent to which policy changes relate to data-driven decision-making based on changing public health risks.

This study examined policy documents of 13 hospitals in Ontario using a digital archive database that captured obstetric policies posted on hospital websites between March 2020 and May 2023. These policies were analyzed to explore three key public health measures with equity implications for perinatal health: visitor policy, care partner restrictions, and doula support. Each of these policy categories were then analyzed from a health equity perspective, which involved identifying which populations may experience unintended health impacts from the policy measures [17]. A thematic content analysis was also performed to highlight impacts on overlooked populations. Lastly, using data from Ontario Public Health, a COVID-19 Risk Assessment index was created for each hospital using public health unit level data from 2020 to 2023 to examine how policy changes reflected data-driven decision-making based on evolving public health risks.

### Conceptual framework

This paper draws on interdisciplinary literature related to social inequities in health, crisis management during public health emergencies, as well as social justice and data-driven decision-making in public health.

#### Health equality vs health equity

Although often framed as a neutral or objective task, public health measures often reflect a lack of clarity between the concepts of equality and equity [8,18]. Health *equality* entails the allocation of the same health services at the same level to all populations [19]. This may be completely ineffective in some situations (as would be the case in the equal distribution of shingles vaccines to all age groups, which would disproportionately benefit the elderly as opposed to children) while it may be unfair in others (such as the equal distribution of COVID-19 vaccines to all social groups, which would leave blue/pink collar workers more vulnerable than white collar workers, who are able to work remotely). As such, processes rooted in equality may be “fair in form, but discriminatory in operation” leading to disparities in health between social groups [20].

On the other hand, health *equity* is normative and value-based, evoking a sense of justice or fairness not only in access to health resources but also in the impact from involuntary exposure to health risks [19,21]. The consensus among scholars is that health equity involves differences in health that are unnecessary, avoidable, and unjust [8,19,22]. Scholars emphasize the need to identify disparities that occur systematically – rather than randomly – between socially advantaged and disadvantaged populations [21,23]. Equally important is the recognition that health is not simply the absence of disease, but rather physical and mental wellbeing [24].

Health equity focuses on avoidable, systemic impacts stemming from differential access to health-protective resources as well as the variation in involuntary exposure to health risks [25]. As such, social determinants of health (e.g., race, gender, employment, education) play a crucial role in understanding what leads to this variation in access and risk and which populations carry a greater health burden [21,23]. Scholars are increasingly calling for greater attention to health inequities experienced by socially disadvantaged populations situated at the intersection of multiple systems of stratification, such as gender, race, and class [25,26].

#### Crisis management & health disparities

In Canada, social and health inequities are well-documented, with studies showing disparities in health outcomes for women, racialized groups, immigrants, LGBTQ populations, individuals with lower levels of education, and people with disabilities [27–31]. Historical events intersect with the underlying social structures that produce these disparities, which serve to further reinforce and exacerbate inequities across groups of Canadians. For example, research on previous pandemics and epidemics has confirmed the magnification of inequalities, with disparities in health outcomes widening during both the 1918 Spanish influenza pandemic and the 2009 H1N1 influenza outbreak [10,32]. Other research on public crises more generally shows inequities in health outcomes between men and women, rural and urban populations, highest and lowest occupational classes, among populations with low educational attainment, among racial/ethnic minorities, and in neighbourhoods with high levels of material or social deprivation [10,32–34]. Researchers have described this amplification of health disparities during public health crises as a *syndemic*, a phenomenon in which disease burden intensifies for disadvantaged populations due to preexisting social inequalities and comorbidities [32,35].

One potential mechanism for widening disparities during periods of crises lies in the intersection of institutional policies, such as hospital policies, and existing health disparities. In 2020, at the start of the COVID-19 pandemic, hospitals implemented policies that restricted or limited the presence of support persons during the intensive process of labour, delivery, and postpartum care [13]. Systematic reviews on the role of birth companions highlight their clinical and emotional importance. Research suggests that birth companions improve the likelihood of spontaneous births; decrease the likelihood of medical interventions, such as C-sections, vacuum, or forceps use; reduce the use of pain medication during labour and the need for labour induction/augmentation through oxytocin; decrease the risk of postpartum depression; and reduce the likelihood of a low APGAR score [36,37]. Furthermore, women who give birth without support are more likely to report negative birth experiences and to experience birth-related depression and post-traumatic stress disorder (PTSD) [38,39] restrictive visitor policies that limit the support available to birthing women may inadvertently result in iatrogenic harm [4].

In addition, some researchers raise the potential for harm of crisis management policies that involve a temporary abandonment of “Mother-Baby Friendly” practices (such as the presence of labour partners and doulas as well as newborn skin-to-skin contact) and a return to a traditional medical model of obstetrics that separated mothers from their infants and families [40]. While some studies have found no significant association between pandemic-era policy changes and adverse maternal or neonatal health outcomes [41,42], others underscore the mental health impacts of these restrictions, which may have intergenerational reverberations [40,43,44]. Moreover, studies report that the mental health of marginalized women in particular may have been disproportionately affected [5,45].

#### Social justice in public health & data-driven decision-making

Scholars argue that public health decision-making and social justice are inherently linked [9,46]. Public health, in principle, serves a dual mandate: to improve population health and to reduce health disparities [47]. Yet, during crises, the second goal is often overshadowed by efforts to protect the majority. Silva and colleagues [9] suggest that despite the best efforts of public health officials, health measures may inadvertently and disproportionately harm disadvantaged populations through three pathways: the oversimplification of evidence, the overreliance on utilitarian ethics, and the underrepresentation of minority voices in policymaking.

These pathways are especially evident in maternal care, where public health responses require a careful balance between safeguarding population health and minimizing harm to marginalized groups. To fulfill these goals, public health officials often draw on past data to inform their responses to emerging health crises and many of these hospital guidelines may have stemmed from practices developed during previous coronavirus outbreaks, such as the SARS (2003) and MERS (2012), which had serious adverse outcomes for mothers and newborns [48]. These experiences likely led clinicians to extrapolate from those cases when faced with the novelty of the COVID-19 virus [49]. Notably, there are other parallels between the SARS epidemic and the COVID-19 pandemic, including scientific uncertainty and an early reliance on isolation for infection control [50]. It is unclear, however, whether policies meaningfully engaged with the dual responsibility to protect population health and reduce health inequities.

Learnings from these public health crises also reflect elements of utilitarian reasoning and centralized decision-making, raising critical epistemological questions such as who decides what constitutes scientific evidence and whose risks count [51–53]. For example, while evidence might suggest an increased risk of viral transmission from support persons during the postpartum period, there is also substantial evidence that postpartum support improves maternal health outcomes, which in turn affects infant outcomes. Importantly, while evidence in 2020 was limited, by 2021 clearer data had emerged regarding COVID-19 transmission, incubation periods, and mitigation strategies, warranting a re-examination of restrictions that affected birthing person and newborns [6,40]. This underscores the need to critically assess how “evidence” is used in public health and analyze how decisions are made during a crisis.

Theoretically, public health policies are developed and implemented by hospital administrators, who operate under a logic of confidence as agents of the state [54,55]. Studies on organizational behaviour during a crisis or threat show a greater centralization of decision-making, an increase in formalization and standardization, and rigidity in procedures [56–58]. This centralization of authority paired with resource constraints may also lead to blind spots and limited diversity in interpretation and sensemaking [59,60]. Decision-makers may also direct attention to internal process efficiencies to regain control over the external threat [58], focus their attention on a limited set of issues [61], and apply contradictory weights depending on biased interpretations of environmental stimuli [62]. Due to information overload, decision-makers may also weigh extreme data points more heavily and rely on cognitive heuristics to simplify decision-making [63–65].

Within the context of COVID-19 pandemic policymaking, these weights may have been derived from data-driven decision-making based on limited information about viral transmission, a focused attention on a limited set of issues, and management of resource constraints. In some jurisdictions, prominent leaders provided daily press briefings using scientific terminology to “flatten the curve” while drawing attention to indicators such as hospital bed scarcity, ICU admissions, and capacity overload [12] This suggests that these indicators likely played a central role in guiding public health decision-making during the COVID-19 pandemic in Ontario.

### Methodology

To examine how maternal care policies evolved during the COVID-19 pandemic across hospitals in Ontario and to investigate the impact of these changes on the health of marginalized populations, maternity-related policies posted on hospital websites between 2020 and 2023 were examined for a sample of 13 hospitals in Ontario. To do this, a digital content archive database (archive.org) was used to identify policy changes made on hospital obstetrics websites. In total, there were 168 policy documents across the 13 hospitals that comprised of a total of 776 pages of hospital policies. Policies posted on websites were selected for analysis because they provide a window into how institutions “represent and account for themselves” and provide an overlooked lens through which to examine the decision-making process of organizations through a public-facing vehicle [66].

The period selected for analysis (between March 2020 and May 2023) represents the beginning and end of the official *emergency* stage of the pandemic, according to the WHO. The choice of time frame was an important consideration, as various governing bodies differed on what they considered to be the end date of the COVID-19 pandemic. This suggests that the “end” of the pandemic was, in fact, a social and political construct. For example, while the Ontario Ministry of Health (MoH) lifted all public health and workplace safety measures by April 2022 [67], the WHO did not declare an end to the COVID-19 public health emergency until May 5, 2023, suggesting that while the emergency phase had passed, the pandemic itself was not over [68].

Of particular interest in this analysis is how individual hospitals chose to interpret and implement a key MoH directive on visitors and essential care (also known as “Directive 2”) by the Chief Medical Officer of Health for Ontario at the time, Dr. David Williams. In this directive, the Office of the Medical Officer of Health “strongly” recommended that acute care settings only allow essential visitors for “a patient who is dying or very ill or a parent/guardian of an ill child or youth, a visitor of a patient undergoing surgery or a woman giving birth” [69]. How administrators and policymakers at each hospital interpreted “essential care” for pregnant, labouring, and postpartum women is especially important in understanding the power and politics of decision-making on an institutional level.

#### Objective 1: Evolving hospital policies

To examine how Ontario hospital maternal care policies evolved during the COVID-19 pandemic, a sample of 13 hospitals was selected from the 93 hospitals in Ontario that provide perinatal, birthing and newborn services [70]. One hospital per public health unit was selected, representing 13 of Ontario’s 34 public health units [71]. While Ontario hospitals do not operate under the authority of local public health units, they coordinate with them on matters of infection prevention and outbreak management [72]. During the COVID-19 pandemic, provincial directives were central, though some hospitals may have adapted policies in response to local public health guidance or transmission patterns. As the Toronto Public Health Unit services a sizable population, a second hospital was reviewed as a consistency check to assess whether policy timing differed substantially within the same health unit. The consistency check showed that policies were implemented at roughly the same time and the comparison did not offer additional insights beyond what was already captured by variation across the 13 hospitals. Thus, this additional hospital was not included in the primary analysis.

Each hospital was selected using the Ontario Ministry of Health published list of hospitals by region (East, West, Central, North, and Toronto) [73]. This study focused on large and medium population centres, since hospitals in these regions were most likely to have a dedicated birthing unit and have the resources to update their policies online more frequently than smaller population centres. For this analysis, each hospital’s website was reviewed using archive.org between March 2020 and May 2023 and then a database was created containing the dates of policy updates, the wording of each policy update, and the researcher’s notes on potential implications for pregnant, labouring, and postpartum mothers. Characteristics of each hospital by city are outlined in the Table 1 below.

**Table 1.**
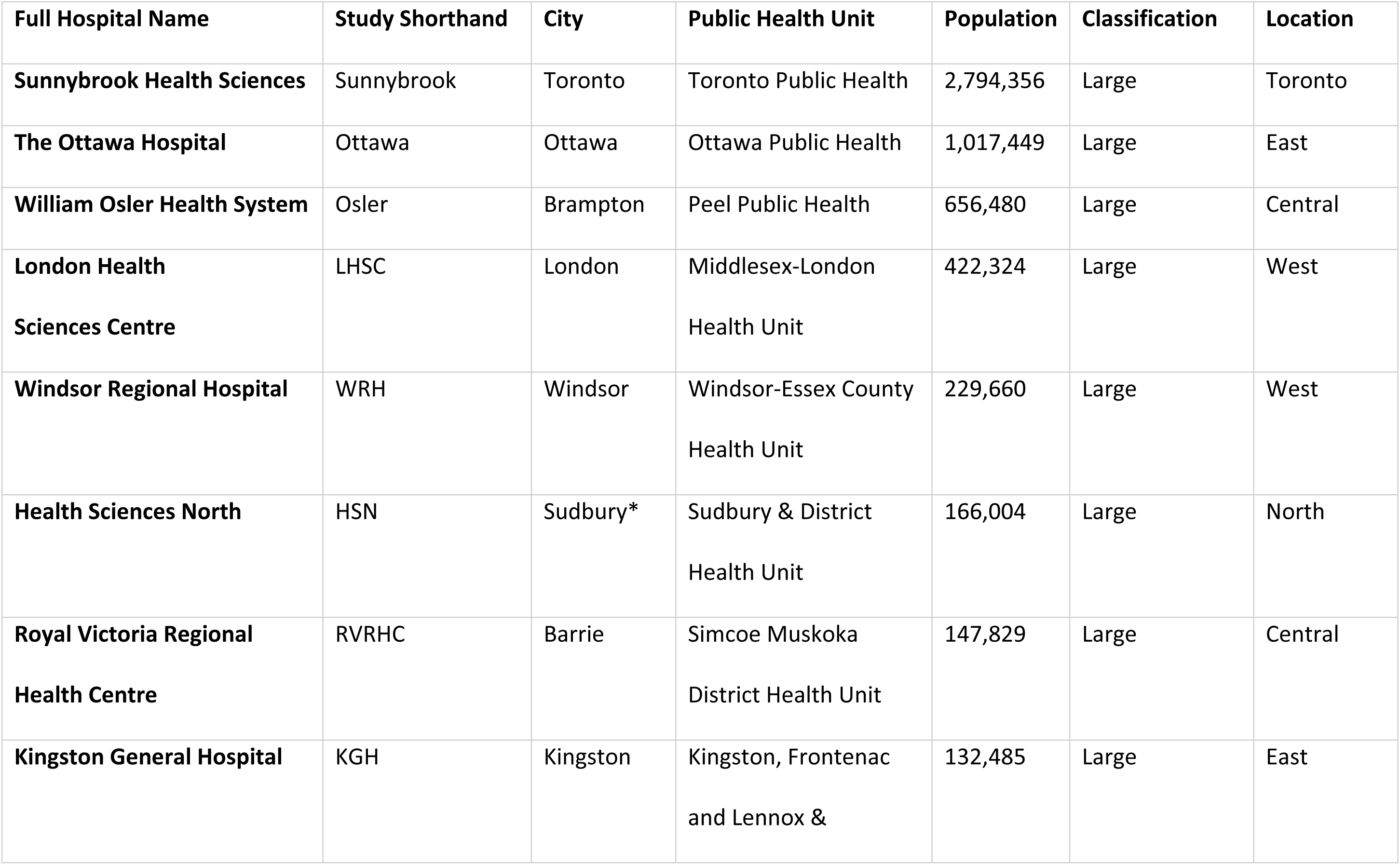
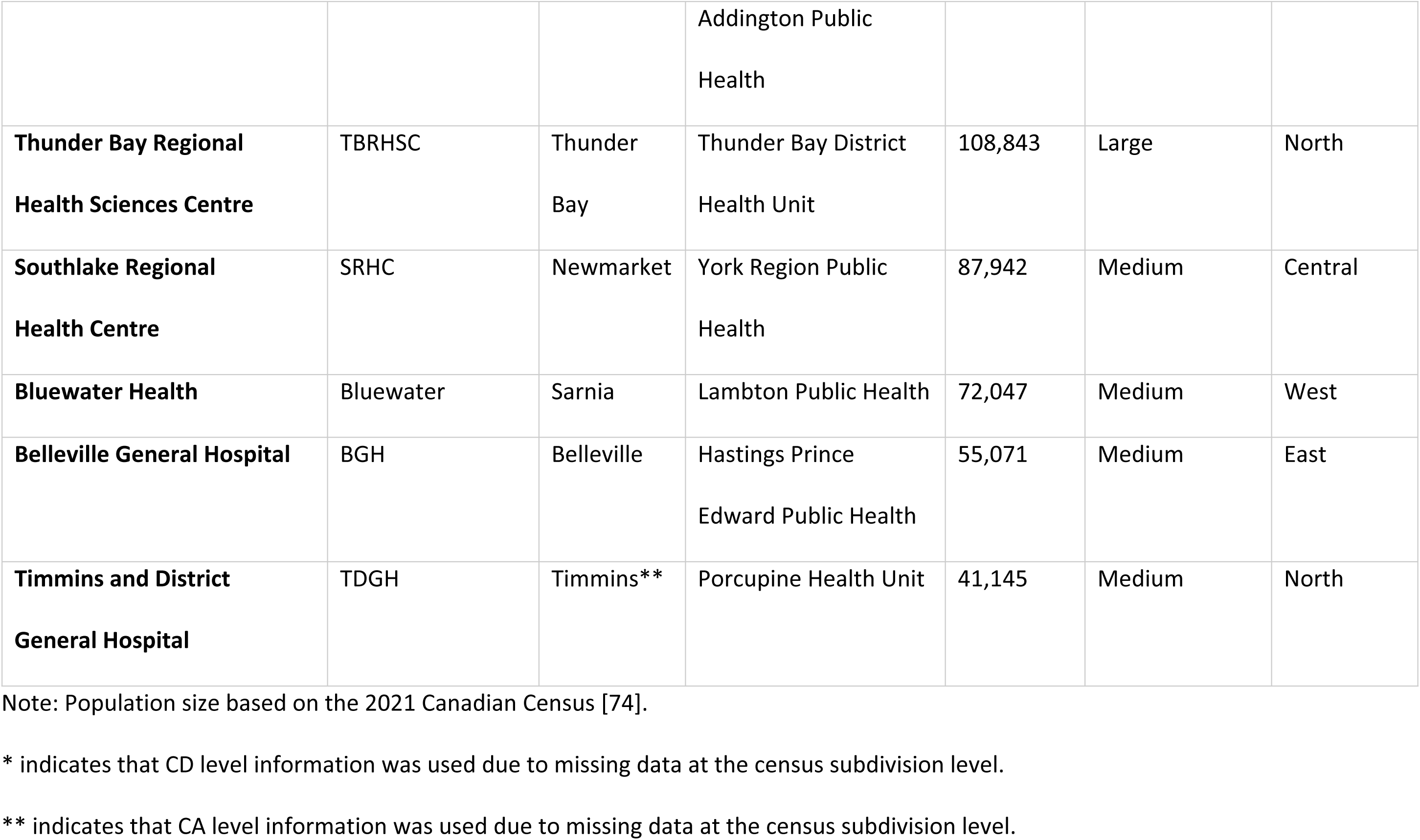
Characteristics of Hospitals Selected for Policy Analysis.

#### Objective 2: Health equity impacts

To address the second objective of this study and assess the health equity impacts of these policies on marginalized populations, the Health Equity Impact Assessment (HEIA) framework (see S1 Appendix) was used. HEIA aids in identifying unintended health impacts from policies and supports in improving policy programs to increase equity-based decision-making [17]. The HEIA framework is centered around a 5-step process: scoping, potential impacts, mitigation, monitoring, and dissemination. For the purposes of this research, the first two steps were used to identify the populations that may have experienced significant unintended impacts because of pandemic birthing policies and determine whether the potential impacts had positive or negative implications for these populations.

For each hospital, the marginalized populations that were most likely to be affected by the policy change were identified. To better understand the heath equity impacts on these marginalized populations, the hospital policy documents were then uploaded into NVivo 15, and a qualitative content analysis was conducted to determine key themes in the policy documents. This methodology was used to reduce the volume of data and employ a systematic approach to identify key themes using a coding frame using a concept-driven and data-driven methodology to identify key codes [75].

**Table 2** shows the coding frame used to determine key themes in hospital policymaking as well as the criteria for inclusion in this study.

**Table 2.** Coding Frame for Thematic Content Analysis of Hospital Policies.

| Parent Code | Subcategory | Meaning | Equity Impacts |
| --- | --- | --- | --- |
| **Essential Support** | Essential Support Partner (ESP); Designated Care Partner (DCP) | Definitions assigned to care partner | Yes, if mothers do not receive essential support |
|  | Postpartum support | Whether partners could offer postpartum support | Yes, if mothers do not receive essential support during this time period |
|  | Prenatal support | Whether partners could offer prenatal support | Yes, if mothers do not receive essential support during this time period |
|  | Triage | Whether partners could offer support during triage | Yes, if mothers do not receive essential support during this time period |
|  | Visitors | Whether visitors were allowed | Yes, if “visitors” is synonymous in policy for essential care partner |
| **Masking** |  |  | Yes, if mother is required to mask during active labour. |
| **Re-entry Restrictions** | Deliveries |  | Yes, if support partners excluded due to essential need to exit hospital |
|  | Food |  | Yes, if support partners excluded due to essential need to exit hospital |
|  | Smoking |  | Yes, if support partners excluded due to essential need to exit hospital |
|  | Leave/Exit |  | Yes, if support partners excluded due to essential need to exit hospital |
| **Vaccination** |  |  | Yes, if mother is left with no support due to vaccine policy. |
| **Testing** | Polymerases Chain Reaction (PCR) test |  | No evident equity implications |
|  | Rapid Antigen test |  | No evident equity implications |
| **Doula** |  |  | Yes, if doulas are excluded as essential care workers |

While all policy categories were examined using NVivo’s keyword search function, not all public-facing documents provided sufficient detail to assess potential equity impacts on marginalized populations. Therefore, this analysis focuses on the three policy areas where there was clear and documented evidence of possible inequitable effects: *essential supports*, *re-entry restrictions*, and *doula care*. These categories were included because the available policy language demonstrated a meaningful potential to differentially impact mothers and families from marginalized groups.

#### Objective 3: Data-driven decision-making

To determine the extent to which policy changes may have been motivated by evolving COVID-19 public health risks, a quarterly COVID-19 Risk Assessment index was created for each hospital using public health unit level data from 2020 to 2023 on three core indicators of system pressure and disease burden (standardized per 100,000 people): COVID-19 deaths, hospital admissions, and hospital bed occupancy [76].

Each indicator was normalized using a min-max method, where values were scaled between 0 and 1 relative to the historical maximum and minimum for that indicator using the following equation [77]:

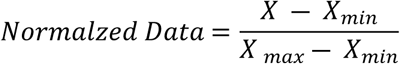

This method ensured comparability across indicators that differed in scale and variance. For each quarter, normalization was based on all historical data available up to and including that quarter (i.e., a cumulative rolling window). This simulates the decision-making horizon of a decision-maker that engages in sensemaking as an ongoing process [59]. Following normalization, a composite index — termed the Operational Burden Model — was calculated by assigning greater weight to indicators of health system strain and over factors which administrators have greater control: 40% to occupancy, 35% to admissions, and 25% to deaths [58]. This model assumes that institutional decision-makers base policy on all available COVID-19 data up to and including the quarter in question, with priority given to indicators of health system pressure indicators. Results from this table are available in S2 Appendix.

To assess the robustness of the findings, two alternative decision-making models were constructed using different indicator weightings. The first, a Rational Choice Model, applied equal weights to all three indicators, reflecting the theoretical assumption that a rational agent would consider each indicator equally when assessing pandemic risk [58]. The second, a Psychological Salience Model (50% deaths, 25% admissions, 25% occupancy), was informed by research on decision-making heuristics, which suggest that managers tend to minimize losses, prioritize recent information, and give greater weight to vivid outcomes such as death [58,64,65]. However, as they produced no substantive differences in the classification of COVID-19 risk levels, only the results from Operation Burden Model are presented. This model aligns with system-pressure based decision-making approaches adopted in other jurisdictions [12], enabling a clearer evaluation of how institutional responses, such as visitor policy changes, corresponded to data-driven assessments of public health risk.

The resulting index reflects each hospital’s relative COVID-19 burden over time, scaled from 0 to 1. For ease of interpretation, scores were then categorized into tertiles (low, medium, high) to classify risk levels across hospitals and quarters. A Gantt chart-based visual was then developed overlaying each hospital’s risk level with its corresponding visitor policy changes as well as population-level vaccination rates, thereby allowing for a visual assessment of whether hospital visitor policies aligned with evolving public health risk.

## Results

This section reviews the policies noted on the sample of hospitals’ websites as related to essential care partners, care partner restrictions, and doula care limitations in Ontario between March 2020 to April 2023. Direct quotes from the hospital’s websites are provided to show evidence of three overarching themes that emerged during this analysis: inconsistency in the conceptualization of essential care in maternal health; policy variability within and between hospitals; and a disconnect between public health risk and hospital policy. Collectively, these patterns suggest an absence of social justice considerations in hospital-level policymaking and point to potential consequences for declining trust in public health messaging.

### Inconsistency in defining “essential care” under crisis conditions

This analysis showed that there was a high degree of variation in hospitals’ understanding of essential care for pregnant people. This lack of a standardized understanding of essential care appears to predate the pandemic, pointing to deeper systemic issues in how maternal health needs are defined and addressed within healthcare systems – and whether psychosocial supports are considered essential in maternal care practices.

As shown in **Fig 1**, the definition of essential care differed greatly across hospitals. While some institutions defined essential care partners for labour and delivery specifically, others offered a more general definition based on MoH directives (specifically “Directive 2” which identified essential visitors only for those “dying or very ill or a parent/guardian of an ill child or youth, a visitor of a patient undergoing surgery or a woman giving birth.”). This grouping of birthing people with surgical patients reflects a medicalized framing of childbirth, emphasizing clinical intervention over continuous psychosocial support. Furthermore, support was narrowly confined to a single point in time – labour and delivery – rather than viewed as necessary throughout the perinatal period. This broad yet medicalized definition gave hospitals considerable discretion in determining what qualified as essential care, and in most cases, policies focused solely on regulating visitor access during labour rather than defining the broader scope of essential birth support.

**Fig 1.**
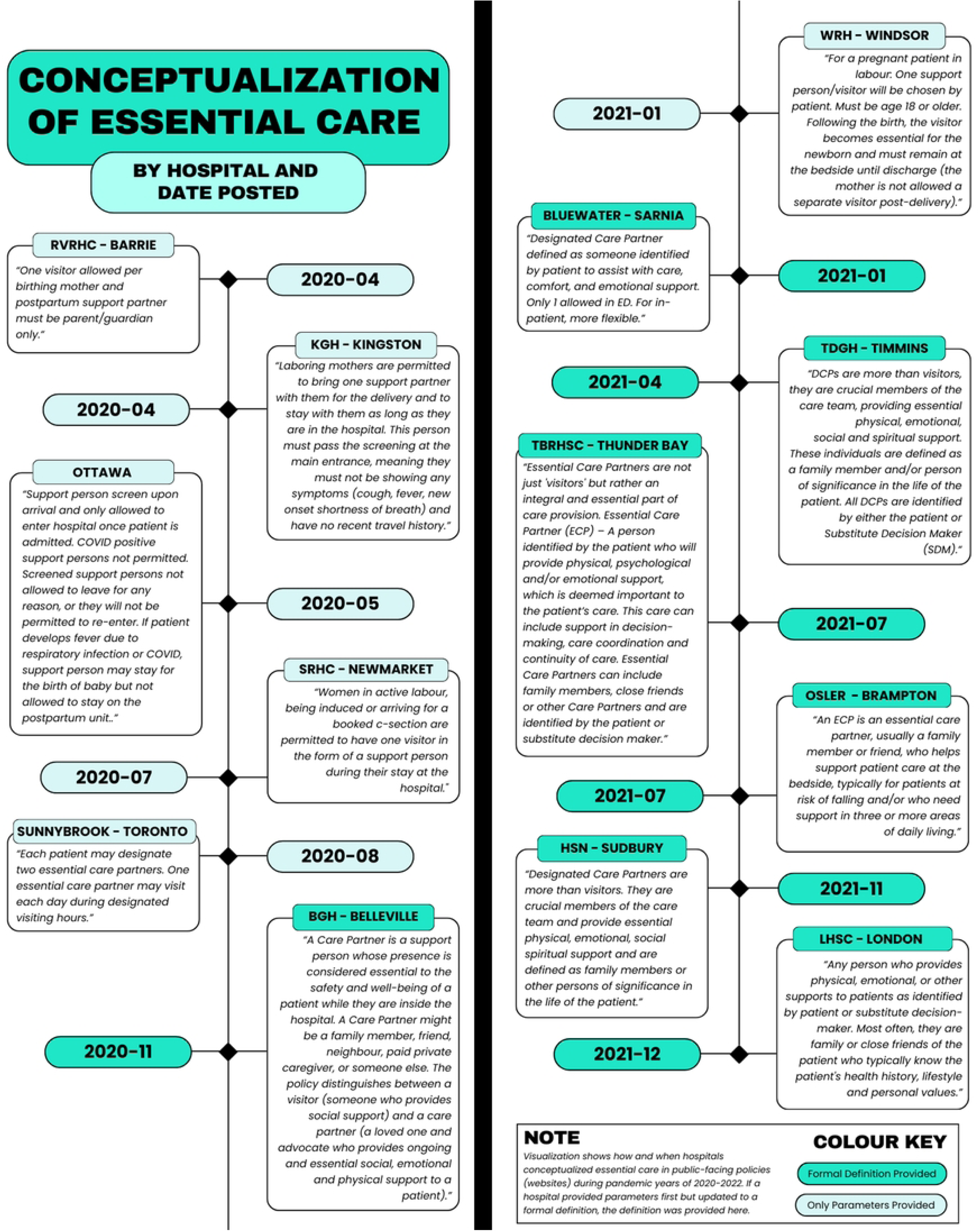
Conceptualization of Essential Care by Hospital and Date of First Update. Visualization shows when hospitals first provided public-facing definitions of “essential care” on hospital websites during the COVID-19 pandemic, highlighting variation across Ontario hospitals and instances where a formal definition was not specified.

One question that arose during the pandemic was whether doulas were considered essential support for perinatal health. In this analysis, most hospitals (12 out of 13) did not include doulas as a part of the medical care team and a little under half (7 out of 13) make no reference to doulas in any of the public-facing documents throughout the pandemic period under study here (2020-2023). One exception was Osler (Brampton), which included doulas as a part of the medical care team as early as April 2021 (the first date of data available for this hospital). Moreover, this policy remained consistent throughout the duration of the pandemic (see Osler in Fig 2).

**Fig 2.**
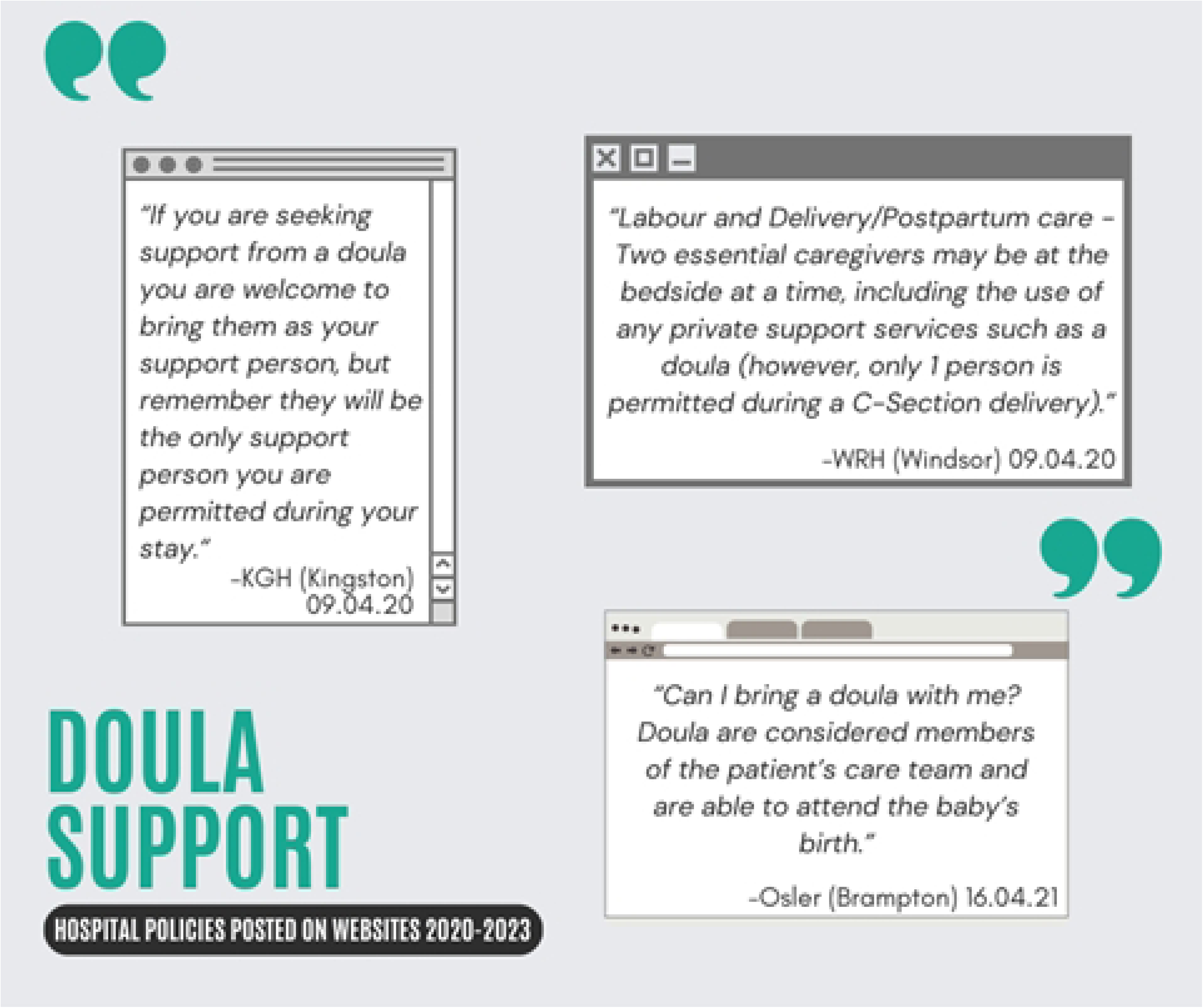
Policies on Doula Support Posted on Ontario Hospital Websites (2020-2023). Visualization illustrates selected hospital policy excerpts regarding doula presence during labour and delivery, highlighting institutional variation in whether doulas were recognized as essential care partners.

As a point of comparison, most hospitals considered doulas to be a support person, not an essential care worker or part of the medical team (see KGH in Fig 2). This can pose an issue for individuals that hire doulas for support for labour and delivery but are then unable to substitute a family member for postpartum care once the doula’s services have been exhausted.

Perhaps in response to local needs, patient advocacy efforts, or lateral learning from other maternity wards, some hospitals began noting their doula policy in 2021, such as Osler (Fig 2) which considered doulas as part of the birth care team. By this stage, some hospitals also began to recognize the need for two support people for a birthing person (see WRH in Fig 2). However, most hospitals did not mention doulas as a form of available care for birthing mothers, and when they did, it was much later in the pandemic. This suggests a shift in perception of doulas as a form of essential support during labour and delivery for women in some hospitals.

In summary, Ontario hospitals varied widely in defining and managing essential support during the pandemic. Most excluded doulas from care teams, and few adopted standardized, evidence-based approaches, raising concerns for equity, continuity of care, and maternal mental health.

### Variability in policy implementation

To address the second objective assessing the health equity impacts of hospital policy changes on marginalized populations, the HEIA framework was used to identify the populations that may have experienced significant unintended impacts because of pandemic birthing policy changes and determine whether the potential impacts had negative implications for these populations. For this analysis, the following policy changes showed evidence of differential impacts on marginalized groups: prenatal and postpartum support restrictions, caregiver re-entry restrictions, and policies that disproportionately harmed minority and vulnerable groups, such as single mothers or women undergoing pregnancy loss. In each section, the marginalized population and associated harms have been identified.

#### Prenatal & postpartum support restrictions

Support during the entire perinatal period (including postpartum) was not uniformly considered a fundamental right of a birthing person. In fact, a support partner was not allowed in the hospital before the mother went into labour in 12 out of 13 hospitals between 2020 and 2021 (at BGH, a support person could stay for ultrasound appointments). This changed in 2021 when LHSC (London) allowed one support partner to accompany a mother being assessed at triage (October 2021), Ottawa allowed one partner for obstetrics/ultrasound appointments (November 2021), and TDGH (Timmins) allowed a partner for ultrasound appointments (December 2021). However, most hospitals had explicit policies that did not allow any support partners during prenatal appointments, including checking into the hospital. For example, Fig 3 shows support partners were restricted from accompanying pregnant persons during the prenatal appointments, ultrasound appointments, and triage prior to admission – a pattern observed across the majority of hospitals in this study.

**Fig 3.**
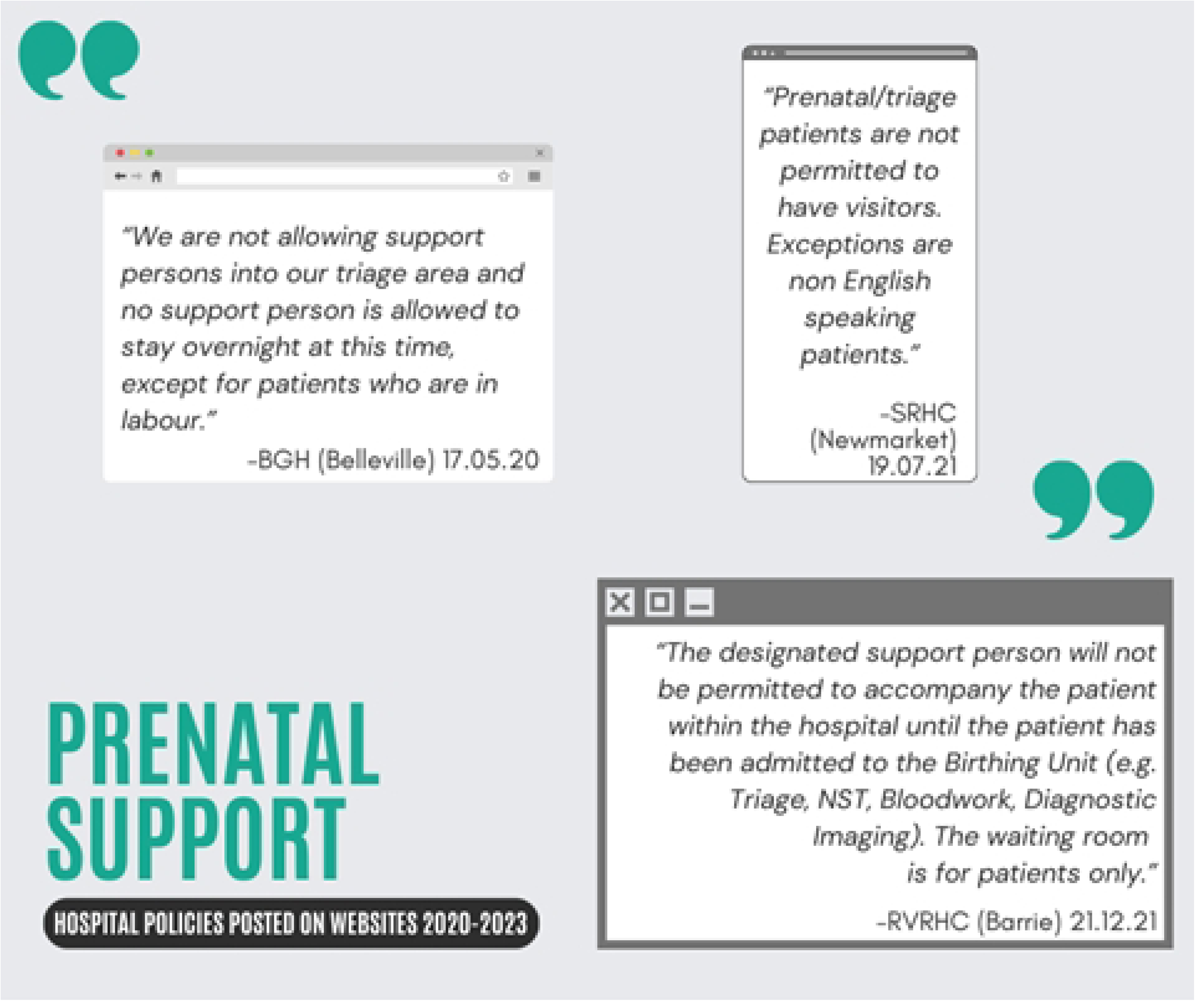
Policies on Prenatal Support Posted on Ontario Hospital Websites (2020-2023). Visualization presents policy excerpts from selected hospitals regarding support person presence during the prenatal period, illustrating restrictions on accompaniment at triage and diagnostic appointments, with limited exceptions.

As a result, many mothers may not have received the emotional support that they needed to ask important questions about their health and the health of their child. In some cases, mothers may have required a partner or support person due to language barriers or difficulty understanding medical terminology. Some hospitals recognized this language barrier and provided an explicit exception later in the pandemic (see SRHC in Fig 3). However, these policies may also have marginalized women with mental health disorders, such as anxiety and prenatal depression, which go largely undiagnosed in women of colour and those with lower levels of education [78].

Aside from labour, many hospitals limited the care available after the birth or did not permit support partners to stay overnight to support the mother (see Fig 4). This policy could have had adverse impacts on first-time mothers, women giving birth via C-section, those with language barriers, and other vulnerable populations. As such, what constituted as essential care for each portion of the perinatal period (prenatal, labour, postpartum) differed greatly across hospitals and may have had adverse health impacts on birthing people, particularly marginalized groups.

**Fig 4.**
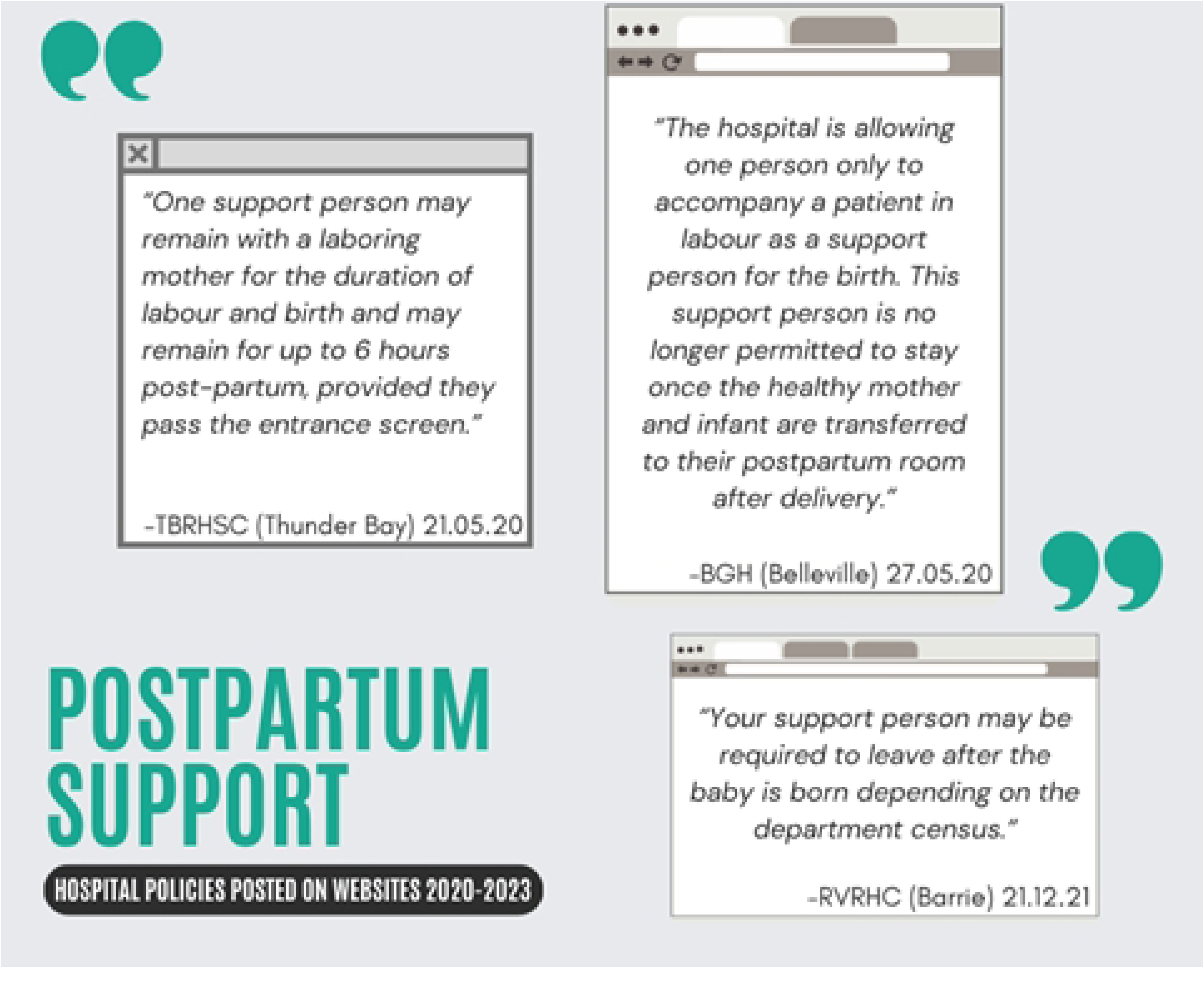
Policies on Postpartum Support Posted on Ontario Hospital Websites (2020-2023). Visualization displays policy excerpts regarding support person presence after delivery, highlighting restrictions on overnight stays and variability in the duration of permitted postpartum support.

#### Caregiver re-entry restrictions

Another area of hospital policymaking with equity implications were re-entry restrictions for support partners, which may have inadvertently contributed to inequitable care partner restrictions. There are at least three essential activities that would have required leaving and re-entering a hospital: to pick up a delivery or obtain essential items (such as food), to smoke (as smoking is not allowed in any public building in Ontario), or to manage other caregiving responsibilities (such as caring for other children).

In this analysis, 7 out of 13 hospitals had explicit policies that prevented care partners from re-entering the hospital if they left for any reason (see Fig 5 for examples of the wording of such policies). Policies restricting re-entry may have prevented people from receiving essential items and affected the quality of labour and postpartum care received. Some hospitals did not allow re-entry for any reason (see Ottawa in Fig 5) while others restricted the number of times a person could leave over a period of time (see Barrie in Fig 5). It is important to note that if these policies were not communicated properly, partners may have missed key milestones, such as the birth of the child. Conversely, there may have been health impacts from such policies if partners had to forgo essential needs to avoid being barred from re-entering the hospital, including access to essentials such as food deliveries (which were not allowed in most of the hospitals in 2020 when data was limited on viral transmission pathways).

**Fig 5.**
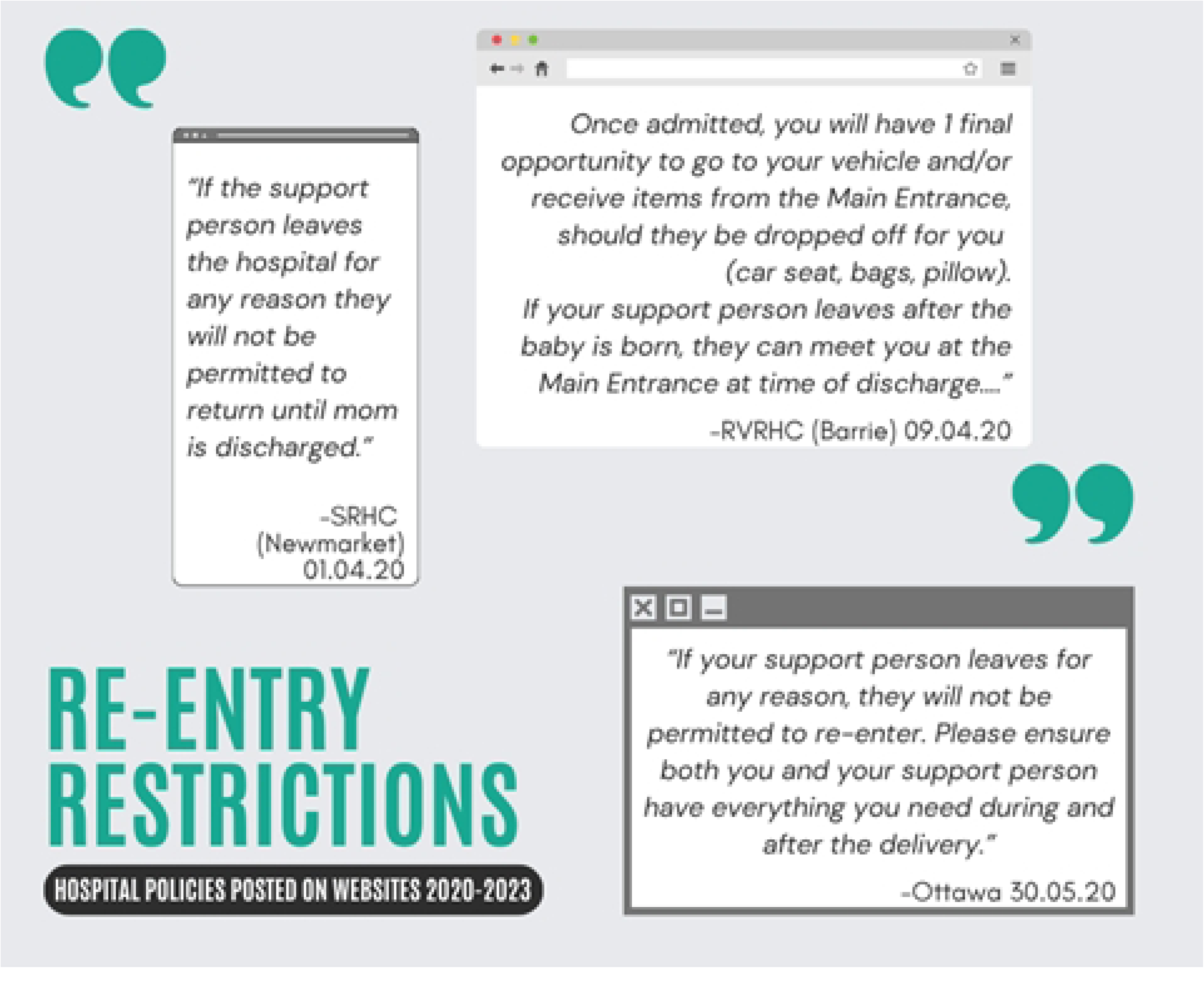
Policies on Re-entry Restrictions Posted on Ontario Hospital Websites (2020-2023). Visualization presents policy excerpts limiting support person re-entry after leaving the hospital, underscoring institutional rules that frequently required continuous presence and restricted access to essential items or caregiving responsibilities outside the hospital.

In-and-out privileges that affected smokers could also have had an impact on the birthing mother if they were unable to cease smoking in advance of the birth (see Fig 6). While smoking cessation during pregnancy is generally encouraged, the phrasing of some policies implies that access to birthing services may be conditional on having quit smoking in advance (see RVRHC in Fig 6). As such, it is unclear what would happen if a mother that smokes is unable to “make arrangements for alternative smoking cessation options prior to admission”.

**Fig 6.**
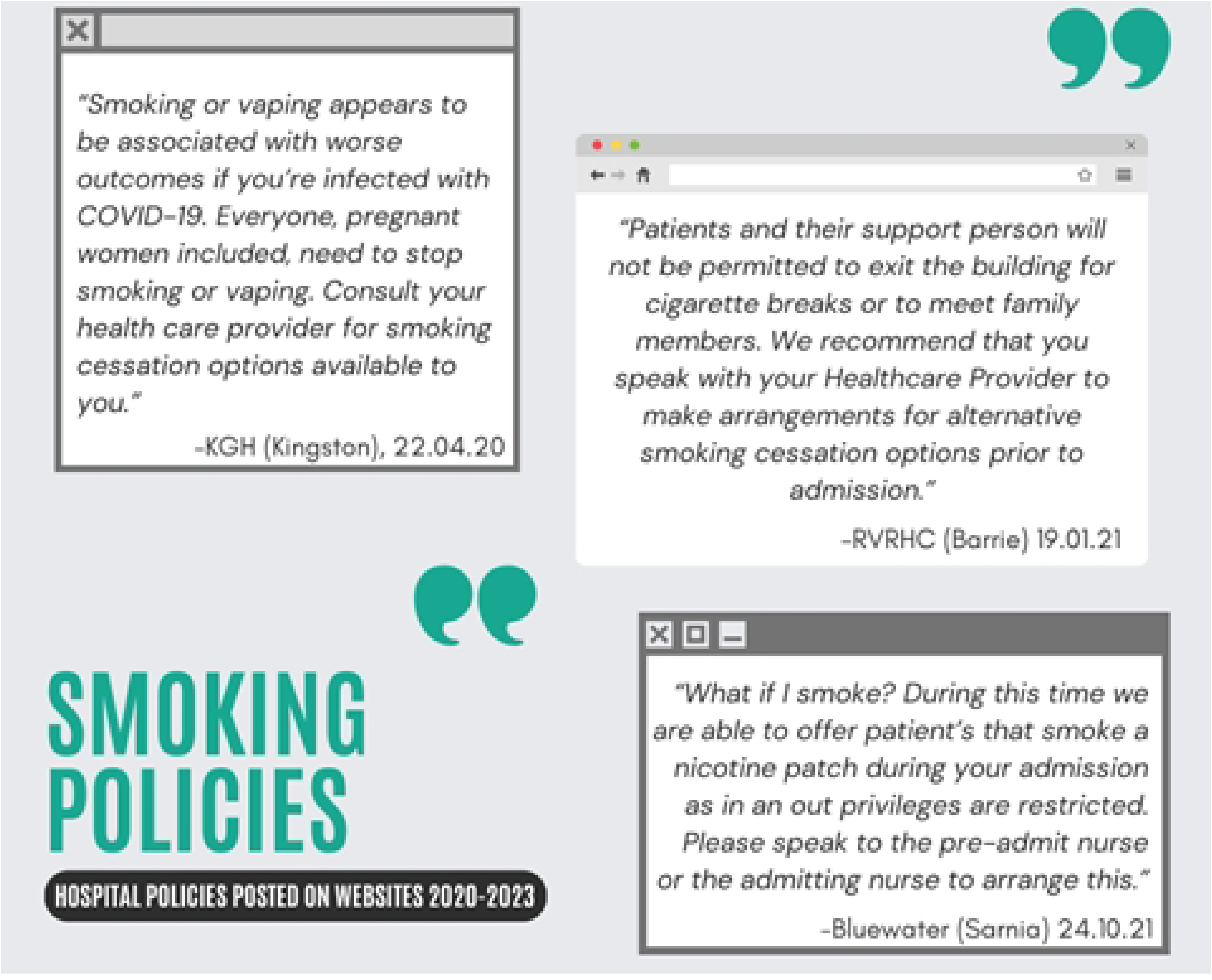
Policies on Smoking Restrictions Posted on Ontario Hospital Websites (2020-2023). Visualization presents policy excerpts addressing smoking during hospitalization, illustrating restrictions on in-and-out privileges for support persons and the expectation of alternative cessation arrangements prior to admission.

It is important to note that framing also matters. In principle, policies that provide justification for the change are most likely to be accepted [64]. One hospital did this effectively by using data and current research to support the policy shift (see KHG in Fig 6). Lastly, while not offered at the onset, one hospital understood the financial cost and time barrier associated with having a healthcare provider arrange this smoking alternative for the patient/visitor (see Bluewater in Fig 6). This universal accommodation method is effective because it allows for use by anyone in need without having to disclose personal circumstances.

#### Other equity implications

There are other equity-deserving groups that may also have been overlooked. For example, policies restricting re-entry or swapping out of care partners (see WRH in Fig 7), may have adversely affected families with other caregiving needs, such as other children. With policies that did not allow the support partner to return after leaving, women with no other social supports may have had to labour alone so their children at home could be looked after by their partner. As such, families with other caregiving needs would have been affected by policies that prevented support persons from leaving the hospital to fulfill other caregiving responsibilities.

**Fig 7.**
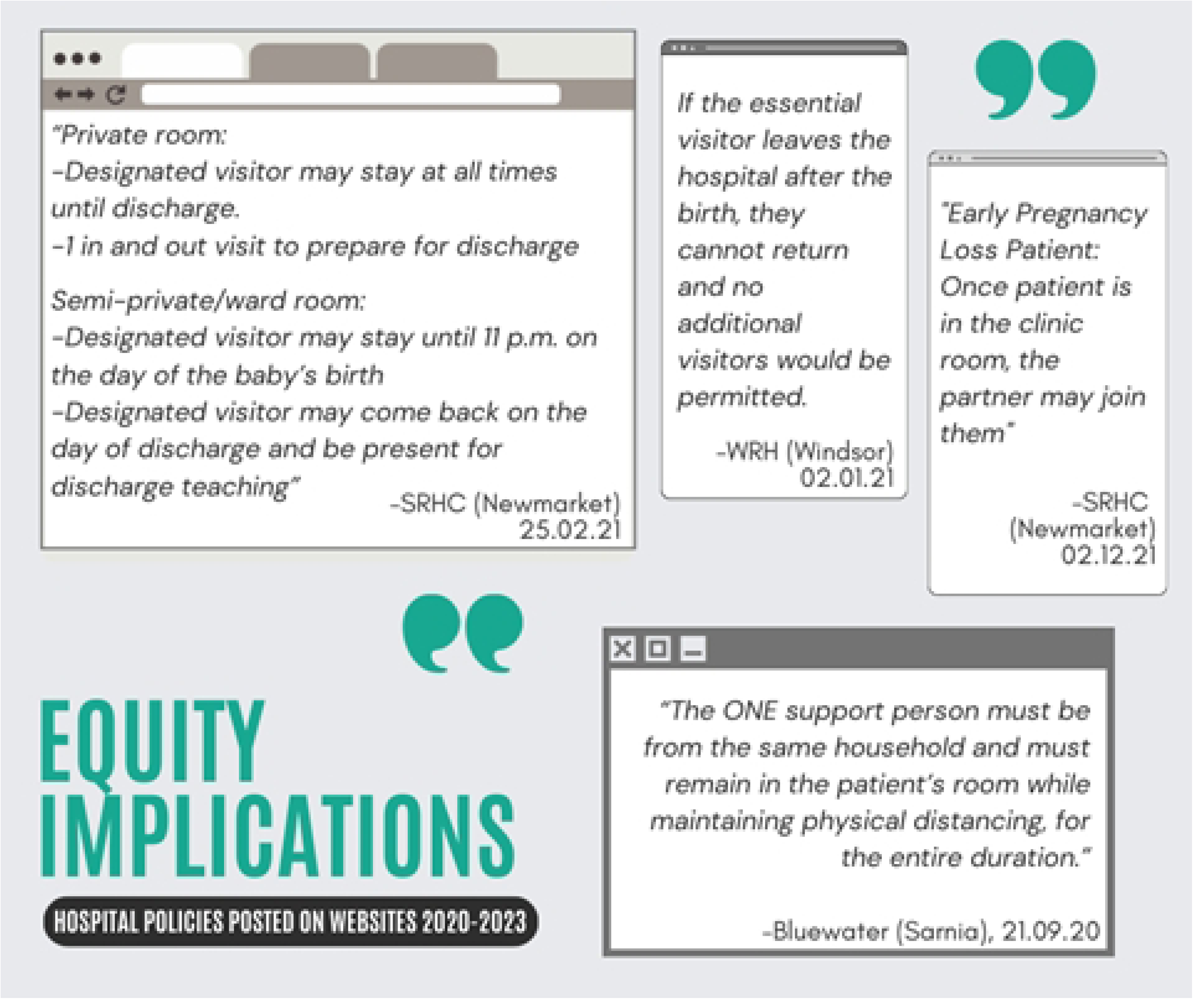
Policies with Equity Implications Posted on Ontario Hospital Websites (2020-2023). Visualization displays policy excerpts that highlight potential equity concerns, including requirements related to household status, pregnancy loss, and private versus shared room access, underscoring how institutional rules could differentially affect families.

Another hospital policy required the support partner to belong to the same household as the labouring mother (see Bluewater in Fig 7), potentially excluding single mothers by choice, such as unpartnered women undergoing fertility treatment, as well as divorced, separated, or widowed mothers. In addition, an overlooked group in policy considerations was women who had experienced miscarriages or early pregnancy loss. In fact, in this study, only 2 out of 13 hospitals make direct reference to this population between 2020 and 2023. Fig 7 shows how SRHC in Newmarket began to include this vulnerable population in policy considerations in December 2021. Lastly, a class divide also emerges when those who can afford private rooms retain access to postpartum support, while others are left without overnight assistance (see SRHC in Fig 7). This has implications for women’s postpartum recovery, especially for those that have undergone C-section surgeries and may require additional support.

In summary, hospitals varied widely in implementing perinatal care policies, often restricting support persons in ways that disproportionately affected marginalized groups. Re-entry bans and the absence of equity considerations for groups such as single mothers, women experiencing pregnancy loss, and those unable to afford private rooms highlight how hospital-level decisions reinforced existing social and class-based inequities.

### Disconnect with public health risks & evidence-based decision-making

To determine the extent to which policy changes may have been motivated and justified by evolving COVID-19 public health risks, a quarterly COVID-19 Risk Assessment index was developed for each hospital and compared against corresponding visitor policy changes and provincial vaccine coverage rates. The findings revealed a misalignment between hospital-level policy decisions and actual public health risk levels. Notably, there appeared to be a lag between the relaxation of hospital restrictions and reductions in public health risk, suggesting that hospital decision-making was shaped more by institutional risk aversion and organizational inertia than by current epidemiological data or vaccine protection levels.

To reduce infection transmission, changing visitor policies became the first line of defence for hospitals. While intended to protect patients and staff, the frequent and inconsistent revisions to these policies throughout the pandemic heightened uncertainty during an already vulnerable phase of childbirth. Across the board, every hospital (except for Brampton, for which there was no data for March 2020), reduced visitors to one person per birthing female at the start of the pandemic, which often led to a difficult choice between having a partner (and often birth parent) present or someone like a mother, sister, friend, or doula who could offer critical emotional and practical support during childbirth and postpartum recovery.

Fig 8 visualizes how hospital policies shifted over time, varied across hospitals, and correlated with the COVID-19 risk levels for that region. Each icon denotes a policy update recorded on the hospital’s website, representing an update to the visitor policy or other COVID-19 measures (e.g., masking, testing, etc.). The type of icon shows whether the hospital policy allowed one birth partner or two at that policy update. The colour for each quarter reflects the COVID-19 risk levels scaled between 0 and 1 relative to the historical maximum and minimum for that indicator (COVID-19 deaths, hospital admissions, or hospital bed occupancy). To contextualize hospital policy response, Ontario COVID-19 vaccine rollout timeline was also included along the bottom of Fig 8, including the vaccination coverage in Ontario for Phases 1 to 3. Ontario’s vaccine rollout progressed in three phases: Phase 1 (Dec 2020-Mar 2021) targeted high-risk groups including long-term care (LTC) residents, healthcare workers, Indigenous populations, and adults over 60; Phase 2 (Apr-Jun 2021) expanded to essential workers, congregate settings, “hot spot” communities, and pregnant individuals; and Phase 3 (May 2021-Sep 2021) opened access to the general public, including adolescents, by which point 75% of the population had received at least one dose and 69% had received two [79,80].

**Fig 8.**
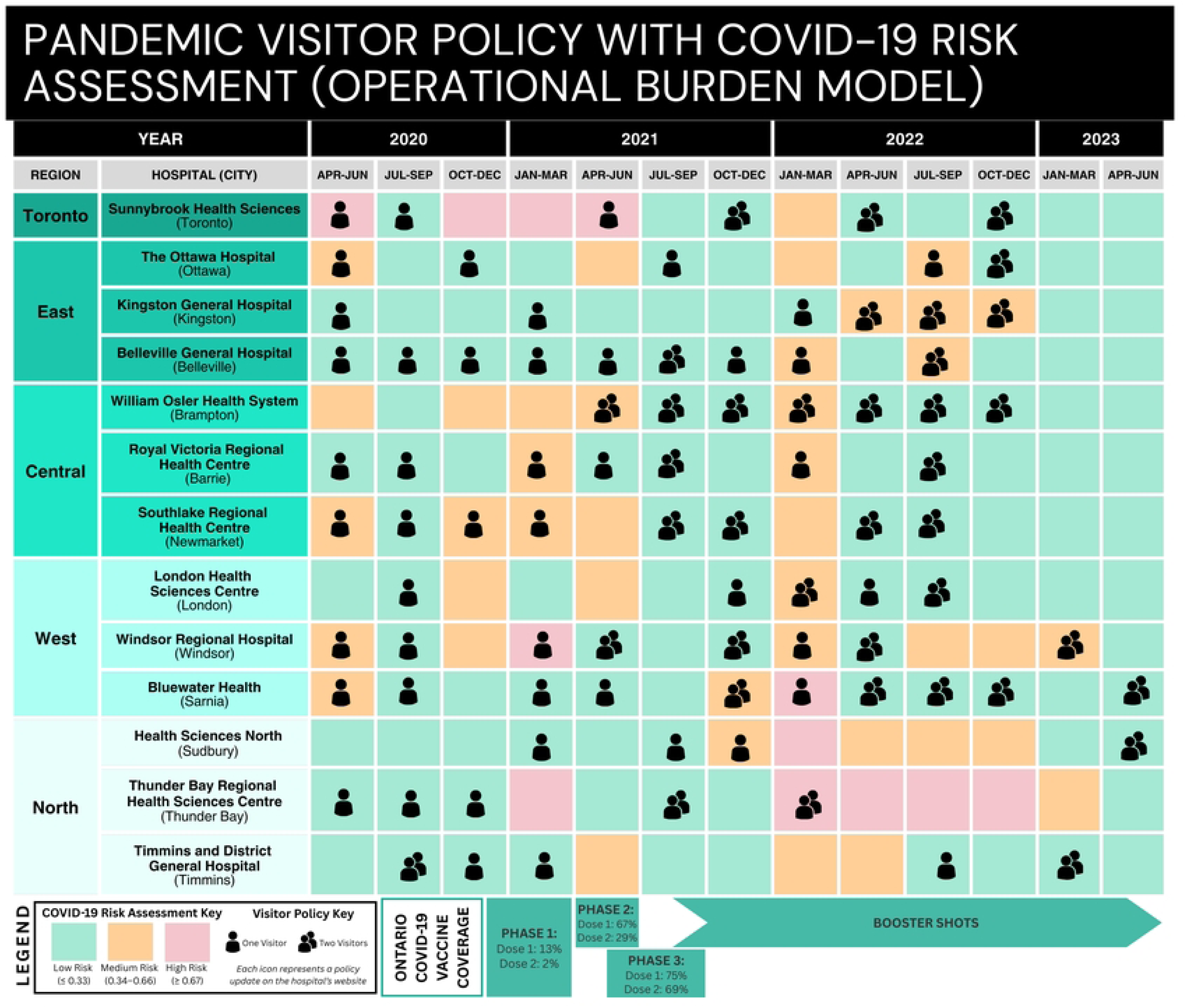
Alignment of COVID-19 Risk Index with Hospital Visitor Policy Updates across Ontario Maternity Wards (2020-2023). The figure presents a quarterly composite COVID-19 Risk Index for each hospital, categorized into tertiles: low risk (green, ≤0.33), medium risk (orange, 0.34-0.66), and high risk (red, ≥0.67). The index is overlaid with visitor policy updates, represented by person icons (one visitor versus two visitors). An icon indicates that a hospital updated any pandemic-related policy (e.g., visitors, testing) during the corresponding quarter. To contextualize policy changes, the visual incorporates Ontario’s COVID-19 vaccine rollout timeline, including vaccination coverage during Phases 1-3.

Notably, visitor policy changes did not consistently align with shifts in COVID-19 risk levels. In some cases, hospitals increased visitor policies to two people even though the COVID-19 risk levels relative to historic data were higher, only to later restrict access again – sometimes within the same quarter (e.g., London, Oct 2021-Dec 2022). In contrast, hospitals like Thunder Bay maintained policies permitting two visitors even amid sustained high-risk periods (Oct 2021-Jun 2022), suggesting a localized interpretation of essential care roles. Interestingly, Osler (Brampton) retained a two-visitor policy (a partner and a doula) across all quarters, regardless of fluctuating risk – possibly reflecting a consistent institutional valuation of doulas as essential care providers.

The vaccine schedule provides further insight into hospital-level policymaking. While 6 out of 13 hospitals expanded visitor access by the end of Phase 3 of the vaccination campaign, others delayed changes until well into the booster campaign, despite widespread vaccine availability and public health messaging indicating strong protection (between 70-80%) from a single dose of the vaccine for up to two months [79]. Moreover, by September 2021, full vaccination was mandated for all staff in hospitals, LTC homes, and community care services, further reducing transmission risk through patient-provider-patient contact chains [6,79]. These delays in policy adjustments, despite declining risk and improved vaccine protection, stand in contrast to MoH guidance and highlight inconsistencies between data-driven public health assessments and actual institutional policy decisions. This may suggest that policies relate more to organizational practices than current context, which aligns with current literature on institutional behaviour and organizational inertia [81,82].

Despite the availability of real-time epidemiological data and rising vaccine coverage, many hospitals were slow to ease restrictions. While some hospitals expanded visitor access in line with vaccination milestones, others delayed changes well into 2022, suggesting decisions were driven more by institutional risk aversion or political signals, such as the official end of the emergency in Ontario, than evidence-based public health indicators.

## Discussion

### Finding 1: Inconsistent hospital policies

This study sought to examine the changes in hospital policies implemented across maternity services during the COVID-19 pandemic with a focus on whether an equity-based lens informed meso-level, institution-specific policymaking, as ascertained through public-facing policy updates on hospital websites. To this end, this study successfully catalogued policies across 13 maternity wards in Ontario between 2020 and 2023. Results showed that hospitals implemented an array of infection control measures, including masking, infection testing, vaccination checks, and visitor screening. Among these, three types of policies were found to have notable equity implications: support person restrictions, re-entry protocol, and doula support limitations.

This study highlights that ambiguous definitions of “essential care” for maternal health at the onset of the pandemic may have shaped policy responses in ways that had a negative impact on the mental – and, consequently, physical – health of pregnant people. As observed in **Fig 1**, only 7 of the 13 hospitals in this study invested time in conceptualizing essential care for a birthing person. This process of defining essential care is not merely administrative; rather, it plays a critical role in protecting maternal and neonatal health. Hospitals that formalized such definitions may have been less likely to withdraw these supports during periods of elevated infection risk, potentially because the removal of defined essential care could be recognized as directly harmful to patient outcomes. A substantial body of research affirms the role of psychosocial supports in improving maternal and neonatal outcomes [36–39], raising the question of why maternal mental health continues to be underprioritized within institutional healthcare frameworks.

Even hospitals that defined essential care did not consistently base their decisions on evidence related to maternal health outcomes. For example, research shows that female support companions improve birth outcomes by shortening labour and reducing interventions [83]; however, policies limiting support to one person forced birthing individuals to choose between a partner and a needed female companion. Furthermore, the exclusion of doulas overlooked the role of trained, paid caregivers – individuals who were prioritized for early vaccination and who posed minimal infection risk – to legitimately assume this role. This pattern reflects a broader neglect of the importance of paid caregiving in providing psychosocial supports and was seen in other settings like LTC and home care settings during the pandemic [84,85]. As illustrated in Fig 8, it is likely that most hospitals did not consider two birthing partners to be essential. By April 2022, two years into the pandemic and after Ontario lifted its state of emergency, only 7 out of 13 hospitals examined allowed two care partners for a birthing person.

While most hospitals understood the importance of at least one support person during labour and delivery, no hospital included the support person in the prenatal stage of the birthing process and policies regarding postpartum presence and duration varied widely. Research suggests that the perinatal timespan is a sensitive period in the life stage of both mother and infant, with lifelong consequences for both groups [86,87] and the removal of birth partners from prenatal appointments may have implications for the mental health of mothers through increased risk of depression [88]. In this study, some hospitals appeared to narrowly focus on labour and delivery as the primary event requiring support, thereby overlooking the importance of prenatal and postpartum care. This is concerning given emerging research identifying the underdiagnosis of prenatal depression [78] and the importance of immediate postpartum period for maternal neurodevelopment and infant bonding [89,90], suggesting these stages warrant sustained attention even during public health crises due to far-reaching implications.

Notably, some hospitals clearly defined essential care in maternity settings and maintained more consistent policies throughout the pandemic (e.g., Brampton). Others extended their understanding of essential care beyond labour to the postpartum period and acknowledged the needs of care partners with greater compassion and flexibility within the realm of evidence-based infection-control measures (e.g., Newmarket). In terms of framing, hospitals that simply outlined the parameters of what a care partner could or could not do appeared rigid and procedural in the policy documents (ex. Barrie or Ottawa). These hospitals also did not update their policy to incorporate other essential services (such as doula care).

An unexpected finding in this study was the emerging recognition of doulas as support partners for birthing people. With the increasing medicalization of childbirth [91], doulas are often excluded from definitions of essential care, as reflected by their lack of Ontario Health Insurance Plan (OHIP) coverage. However, a body of literature is emerging on the importance of these professionals in improving birth outcomes, especially for marginalized communities such as racialized and migrant populations [92,93]. While unclear through the policy documents, advocacy efforts or patient demand may have propelled doulas to be considered essential care. By the end of the pandemic, 8 out of 13 hospitals had made some reference to doulas as a part of the care team. This shows a growing legitimization of doulas as maternal health professionals and essential support partners for birthing people.

### Finding 2: Variation in health equity impacts

This study also identified several populations likely to be disproportionately affected by hospital restrictions, including first-time mothers, single mothers, immigrant or racialized individuals more reliant on psychosocial supports, women experiencing pregnancy loss, those with language barriers, as well as women requiring support following health interventions such as C-section surgeries. The study also documented periods of reduced care that may have negatively impacted mental health, particularly during the prenatal and postpartum periods. Restrictions on support persons also had potential equity impacts, limiting their ability to care for other children, access essentials like food, or navigate health behaviours such as smoking.

Findings show a high degree of variability in policymaking both *within* each hospital over time and *across* hospitals in Ontario during the pandemic. At the hospital level, policies were implemented or rescinded at least 85 times over 2.5 years across the 13 hospitals in this study (the actual number may be higher, but this study was limited to the policy versions captured by the digital archive database). Populations with limited digital literacy or official language fluency may have especially been susceptible to confusion or misunderstandings stemming from the frequent policy changes. This variation may have also affected trust in public health measures with studies showing that rapidly changing policies can erode public trust [94,95] and a breakdown in institutional trust can further affect compliance with future health directives [96]. As such, thoughtful, consistent policymaking for essential care may play a broader role in maintaining trust and have downstream effects on vaccine uptake, lockdown compliance, and transmission prevention.

In addition to variation within each hospital over time, there was also considerable variation between hospitals, highlighting inconsistencies in how provincial guidance was implemented and revealing another layer of inequity: regional disparity. In the US, regional differences in maternal health are well-documented and one study found that 35% of the variation in severe maternal morbidity among racialized populations was attributable to the delivery hospital [5]. This study provides evidence that women in Ontario received different levels of psychosocial support depending on the hospital and region in which they delivered. For example, women delivering in Brampton had access to both a partner and doula throughout the pandemic, while women delivering in Newmarket received meals for themselves and their partner directly from the hospital. Similarly, hospitals like London and Kingston used more compassionate language through framing their policies, which may have helped reduce anxiety during an already unpredictable time.

This highlights the need to examine the high degree of loose coupling; that is, the disconnect between centralized MoH mandates and their varied implementation across hospitals [97]. This study provides evidence of inconsistent implementation of ministry directives, which may stem from limited oversight from a centralized authority and non-transparent hospital-level decision-making (often occurring behind closed doors and with limited input from those most affected). A formalized, province-wide definition of essential care, specifically within the context of maternal care, could mitigate this variability since the removal of *essential* care would equate to mental or physical harm. As such, centralized bodies, such as the MoH, should formally define essential care through consultation with experts-by-lived experience and marginalized groups, thus enabling more consistency within and between hospitals and advancing public health’s broader goal of social justice.

### Finding 3: Unclear logics for decision-making

Finally, this study evaluated whether hospital policy changes reflected data-driven decision-making and aligned with public health risk by overlaying visitor policy updates with the corresponding quarterly COVID-19 risk index for each region and vaccine campaign and coverage for Ontario. Results showed that hospitals were inconsistent in applying health measures and had significant discretion in when and how to apply public health mandates. Results highlight the misalignment between hospital policy updates and indicators of COVID-19 severity (including population-level vaccination progress). This is not unexpected given extensive literature that underscores healthcare infrastructures as highly bureaucratic and rationalized [81,98] as well as slow, inflexible and obstinate to change [82,99].

Moreover, while this analysis shows evidence of restrictive health measures to protect the population at large, it remains unclear whether equity considerations guided hospital-level decision-making, as many policies remained unchanged despite declining disease burden and widespread vaccination coverage. Literature on previous public health crises shows how policymakers often make decisions that neglect equity in policymaking, which can have unintended health consequences for marginalized populations [10,32,100]. This may have also occurred during the COVID-19 pandemic for vulnerable women and their families.

While visitor and support restrictions were implemented to limit infection spread, the result was the potential loss of essential care, especially for vulnerable groups that rely social supports to compensate for structural inequities. Policies such as those limiting birthing support, may inadvertently impact marginalized groups, who tend to rely on social supports to compensate for a shortage in other resources [101–103]. Within maternal care specifically, research shows that minority women often rely on social support systems not only to meet healthcare needs, but also to navigate unfamiliar care environments [104,105]. In fact, one reflection of the importance of psychosocial care would have been to include support persons in vaccine eligibility alongside pregnant individuals. Even as vaccines became available to pregnant people by April 2021, their support partners were excluded, reflecting the broader medical system’s tendency to overlook the role of support persons in maternal health.

The combination of high variability in policymaking and a perceived lack of evidence-based decision-making may have contributed to disenfranchisement and institutional mistrust. For example, the extent to which women opted out of hospital births entirely, choosing to instead give birth at home (through midwifery support) to bypass the stringent and contradictory policies within hospitals is an important empirical question. This has the potential to reinforce inequalities, with only women with previously good health or healthy pregnancies able to opt out of the hospital system. Moreover, this loss in trust in public health systems may have far-reaching consequences in other spheres, such as vaccine uptake, healthcare utilization, or fertility intentions [106].

Taken together, these findings offer insights into how institutional policies shaped birthing experiences during the COVID-19 pandemic by highlighting the potential impacts on health equity and trust in public health and addressing whether public health measures serve to both safeguard population health while reducing health disparities in times of crises. As future public health crises emerge, it is critical that institutional decision-making is grounded in a holistic understanding of patient needs, especially for marginalized communities that may rely more heavily on psychosocial supports as coping mechanisms in times of stress and uncertainty.

## Limitations & conclusion

This study has a few key limitations. First, the availability of data was limited to the frequency with which hospital websites were captured through archive.org, which means that there may be pockets of time during which policy changes occurred but were not captured in this study. Second, there may be a disconnect between policies that were posted on hospital websites and those actually implemented in-person. As such, this study has no way to determine if front-line workers bypassed centralized directives from hospital administration for the sake of more compassionate in-person care. This study also focused on medium and large hospitals with accessible digital records and may not reflect the experiences of smaller or more rural institutions with fewer online updates. Lastly, there may have been lags between policy updates and actual website updates, which shows the disconnect that may exist across other hospital functions.

Hindsight is 20/20 and this study was an exercise in considering equity and social justice in public health mandates. A key takeaway is the involvement of experts-by-lived-experience, such as birthing people, in defining what constitutes as essential care. The time invested in determining what is considered essential support may well ensure more equitable health outcomes for marginalized populations. Similarly, the involvement of professional bodies, such as those representing doulas, when developing policies may help to provide important perspectives counteracting the dominant medicalized model of birth and improving equity-based decision-making in healthcare. Finally, clearer centralized guidance, transparent communication, as well as consistent and compassionate framing are essential to rebuilding trust in public health measures in future emergencies.

## Data Availability

All data used in this study are publicly available. Hospital policy documents were accessed through each institution’s publicly available website, archived and reviewed using digital snapshots between 2020 and 2023 using archive.org. COVID-19 epidemiological data for the risk assessment index were obtained from Public Health Ontario, accessible at: https://www.publichealthontario.ca/en/Data-and-Analysis/Infectious-Disease/Respiratory-Virus-Tool.

## Acknowledgements

I would like to thank my doctoral supervisor, Dr. Kim Shuey, and committee member, Dr. Tracey Adams, for their valuable feedback and guidance on this manuscript.

## Supporting information

**S1 File. Health Equity Impact Assessment (HEIA) framework.** This file provides the HEIA framework used to guide this study.

**S1 Table. COVID-19 Risk Assessment.** This file provides the risk assessment model output tables.

